# Machine learning prediction algorithms for 2- , 5- and 10-year risk of Alzheimer’s, Parkinson’s and dementia at age 65: a study using medical records from France and the UK General Practitioners

**DOI:** 10.1101/2025.01.22.25320969

**Authors:** Thomas Nedelec, Karim Zaidi, Charlotte Montaud, Octave Guinebretiere, Pyry Sipilä, Dang Wei, Fen Yang, Anna Freydenzon, Antoine Belloir, Nemo Fournier, Nadine Hamieh, Beranger Lekens, Yanis Slaouti, Allan McRae, Baptiste Couvy-Duchesne, Yulin Hswen, Fang Fang, Mika Kivimäki, Manon Ansart, Stanley Durrleman

## Abstract

**Background:** Leveraging machine learning on electronic health records offers a promising method for early identification of individuals at risk for dementia and neurodegenerative diseases. Current risk algorithms heavily rely on age, highlighting the need for alternative models with strong predictive power, especially at age 65, a crucial time for early screening and prevention.

**Methods:** This prospective study analyzed electronic health records (EHR) from 76,427 adults (age 65, 52.1% women) using the THIN database. A general risk algorithm for Alzheimer’s disease, Parkinson’s disease, and dementia was developed using machine learning to select predictors from diagnoses, and medications.

**Results:** Medications (e.g., laxatives, urological drugs, antidepressants), along with sex, BMI, and comorbidities, were key predictors. The algorithm achieved a 38.4% detection rate at a 5% false-positive rate for 2-year dementia prediction.

**Conclusion:** The validated prediction algorithms, easy to implement in primary care, identify high-risk 65-year-olds using medication records. Further refinement and broader validation are needed.

## Background

Neurodegenerative diseases (NDDs) represent a major public health challenge in Western societies and one of the greatest drug development challenges. Although there are currently no curative treatments for the majority of NDDs, management of cardiometabolic risks and lifestyle intervention may delay the onset and enhance patients’ quality of life. An essential element in implementing timely primary and secondary prevention measures involves identifying at-risk patients, ideally at the general practitioner settings, well before the onset of the disease. In contrast to vascular diseases, where cost-effective and non-invasive methods such as cardiac ultrasound or supra-aortic trunk examinations are employed, performing of MRI or PET scans, or lumbar punctures to assess risk of NDDs for the total population aged 65 or older is not feasible, ethical or cost-effective.

To enable targeted prevention, various multifactorial risk prediction models have been developed to distinguish individuals at a high risk of dementia, who may benefit most from preventive measures, from those at low risk. Widely used prediction models in research and clinical trials include Cardiovascular Risk Factors, Ageing and Dementia [CAIDE-Clinical],^2^ CAIDE supplemented with APOE status,^3^ Brief Dementia Screening Indicator [BDSI],^4^ and Australian National University Alzheimer Disease Risk Index [ANU-ADRI].^5^ However, recent findings from a study on the UK Biobank revealed that these models missed between 84% to 91% of participants with incident dementia when calibrated to achieve a 5% false-positive rate.^6^ Thus, current predictive models are not capable of identifying people at high risk of dementia at a given age (e.g. 65) when screening of risk groups for chronic conditions is typically implemented.

One approach to improve cost-effective prediction of NDDs is to develop and validate risk algorithms for same-aged individuals by leveraging machine learning on electronic health records. Successful use of machine learning to develop predictive models has already been reported for the risk assessment of pancreatic cancer,^7^ and for the QRISK score,^8^ a cardiovascular disease prediction risk score that complements the established Framingham risk score. Concerning dementia and specific neurodegenerative diseases, several papers have attempted to predict dementia including Alzheimer’s disease (AD) ^9,10^ and Parkinson’s disease (PD) ^11^ using electronic health records, but these efforts have been restricted to a single country and incorporating age as a primary feature in the model. This approach presents challenges in interpreting the Area Under the Curve (AUC), as age often drives the model.

To address this issue, we propose a framework for estimating the risk of dementia at specific age. Drawing inspiration from the French Minister of Health’s initiative to reimburse preventive visits at age 65, with a specific emphasis on NDDs prevention, we aim to develop and validate algorithms estimating the risk of AD, PD, and dementia at this critical age. By harnessing readily available electronic medication data, accessible to general practitioners (GPs), we identified medication patterns and health conditions associated with an elevated risk of AD, PD and dementia and developed algorithms that can be derived without laboratory measurements and easily integrated into primary care settings for efficient risk assessment by GPs. These algorithms could serve as a pre-screening tool, identifying a subgroup of individuals eligible for further, more costly diagnostic tests. This includes regular administration of the Mini-Mental State Examination (MMSE) during consultations or referrals to memory clinics for comprehensive evaluations.^12^

## Methods

### 2.1 Data

We investigated health conditions and prescription of drugs associated with subsequent AD, PD and dementia in France and the UK using the THIN database.^13^ THIN is a large standardized European database of non-extrapolated electronic health records collected at the physician level. The THIN database contains medical records, prescriptions, and diagnoses from 2,500 GPs in France and 532 general practices in the UK and represents 8 million patients in total.^14^ The UK database, accounting for around 6% of the UK population, is representative of the UK general practice population in terms of demographics and type of consultation.^16–18^ The French cohort is representative of the French general population in terms of age, sex, and residence.^15^ For each patient and visit, the diagnosis and prescription established during the visit, as well as all ongoing prescriptions and associated diagnoses, are available. Information in the database also includes records of health indicators, such as weight and height.

### 2.2 Study Participants

For both UK and French THIN cohorts, we extracted all patients born in 1940 and 1945 (aged 65 and 70 in 2010). Patients must be active, i.e. followed by the general practitioner, in 2008, the start of the exposure period from January 1^st^, 2008, to December 31^st^, 2010 (Figure 1). We excluded patients with no diagnostic records before December 31^st^, 2010, or with no diagnostic records after 2011 (eFigure1), or patients who became inactive, i.e. not followed by the general practitioner anymore or died before 2011. Patients who had already been diagnosed with one of the three outcomes before 2011 or with undefined sex were additionally excluded (for a flow chart, see eFigure1 in supplementary materials).

**Figure 1:**
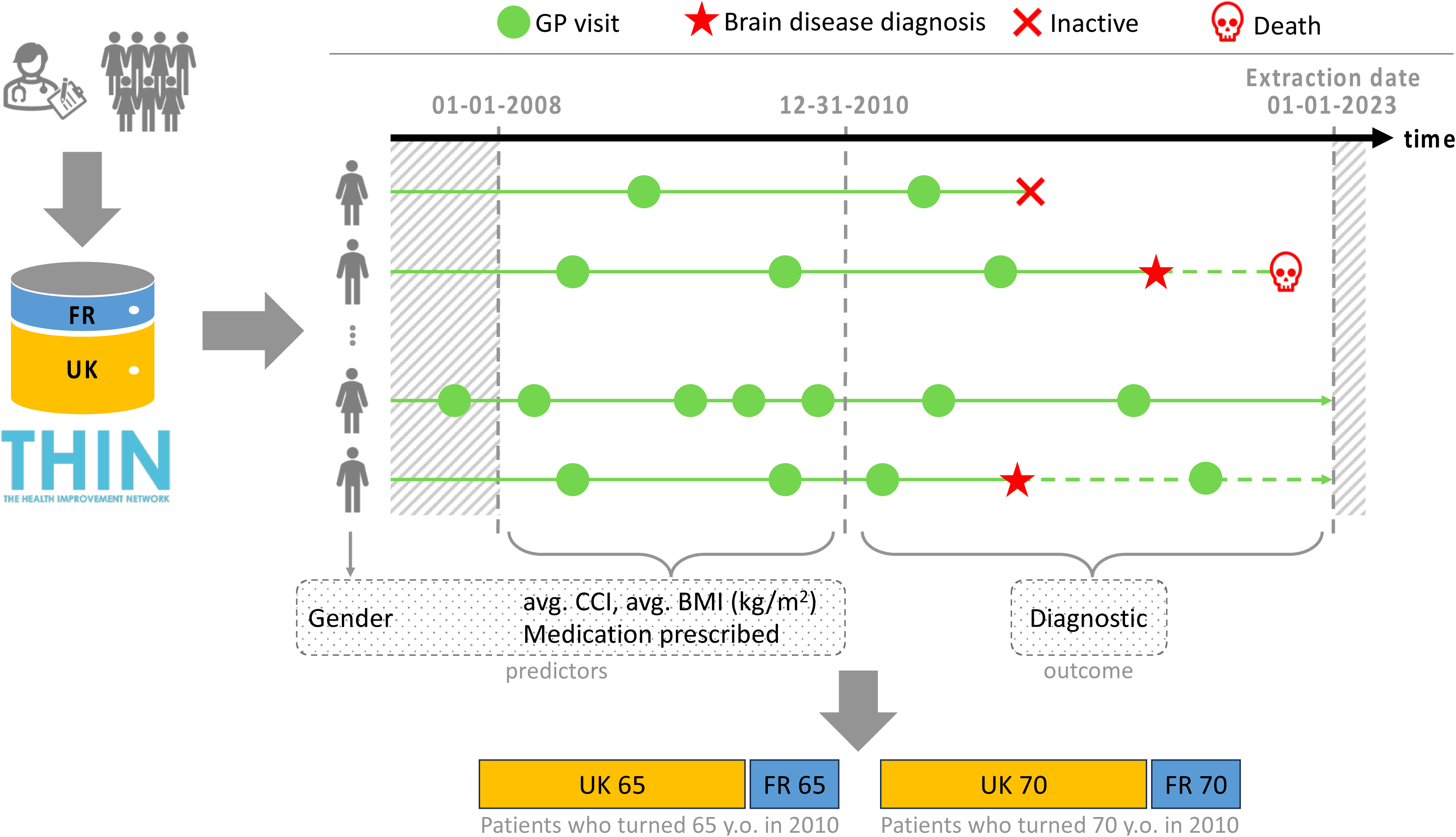
Summary of study design: Extraction of medical prescriptions between 2008 and 2010 for patients who were either 65 or 70 years old in 2010 in France and UK

### 2.3 Outcome

The outcomes of interest were AD, PD, and all-cause dementia occurring between January 1^st^, 2011 and January 1^st^, 2023. We defined the outcome according to diagnostic codes in the International Classification of Diseases, 10^th^ Revision (ICD-10) which were provided directly in the French database, with the exact list of codes available in the supplementary materials. For the UK database, we converted Read codes into ICD10 codes according to the correspondence provided by the UK Clinical Terminology Centre.^16^

### 2.4 Predictors

Exposure parameters included BMI, Charlson Comorbidity Index (CCI), prescribed medications (categorized using the first 3 characters of the ATC code), and medical diagnoses (categorized by ICD-10 codes). To select medications as exposure features for our model, we employed a Fine and Gray survival model to address the competing risk of death. We analyzed patients born in 1940 and 1945 together. Drug families (determined by the first 3 characters of the ATC codes) with a prevalence of at least 5% in the UK cohort were investigated for their association with the onset of the three diseases of interest (Figure 2). We computed Hazard Ratios (HRs) for each medication’s association with Alzheimer’s disease (AD), Parkinson’s disease (PD), and dementia using the Fine and Gray model, adjusted for sex, BMI, CCI, and age group (Figure 3, panel B and eTable 1). Multiple comparisons were addressed using Bonferroni correction (N = 43). We then recalculated HRs in a multivariable model that included all medications associated with the outcomes in the univariate analysis, along with the adjustment variables. To identify the precise medication subclasses responsible for the associations, we performed a more detailed analysis of the ATC hierarchy, exploring two additional levels (first 5 characters of the ATC codes) for medications with a minimum 5% prevalence (Figure 3, panel A).

**Figure 2:**
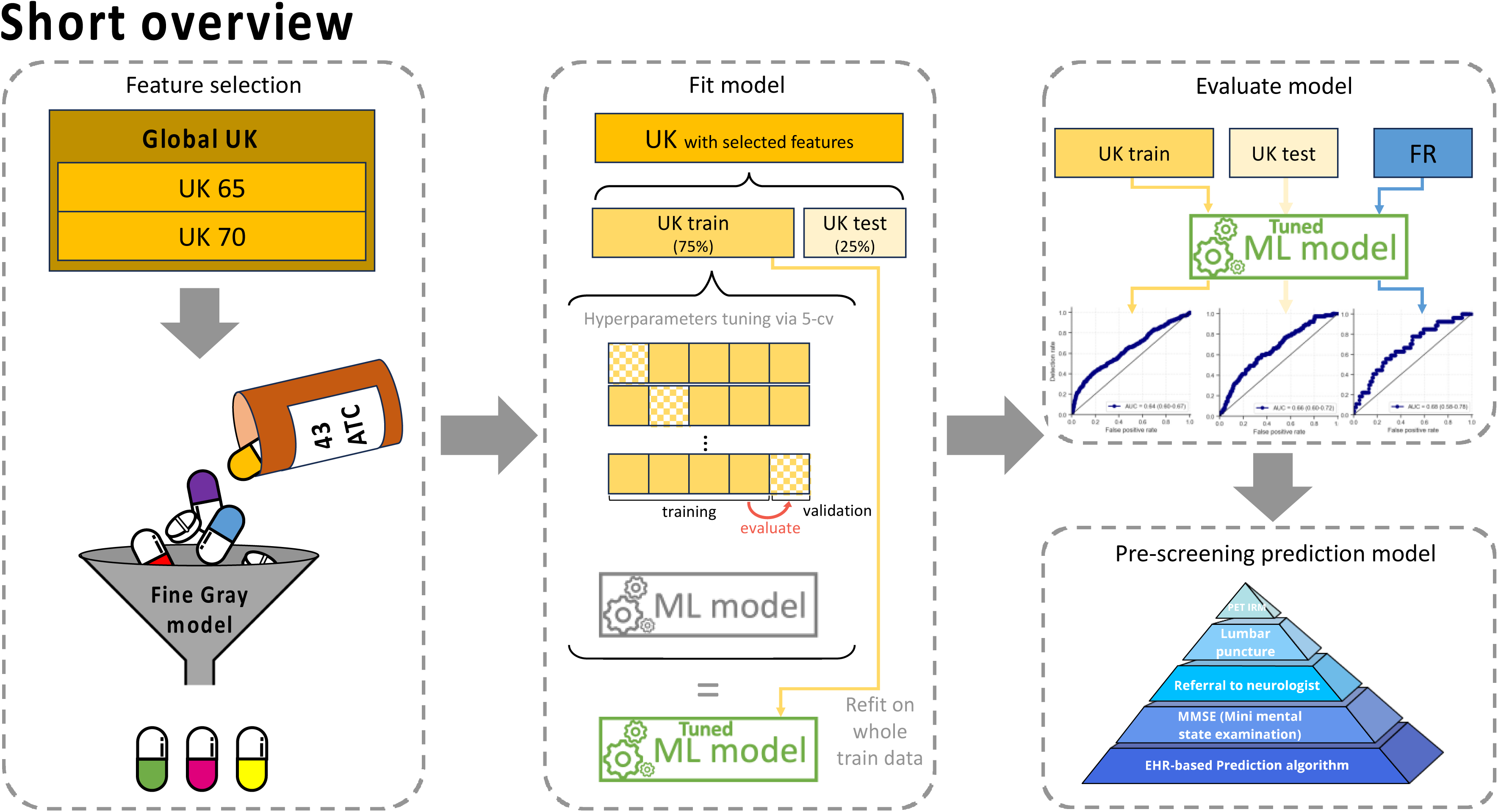
Overview of the modeling strategy

**Figure 3:**
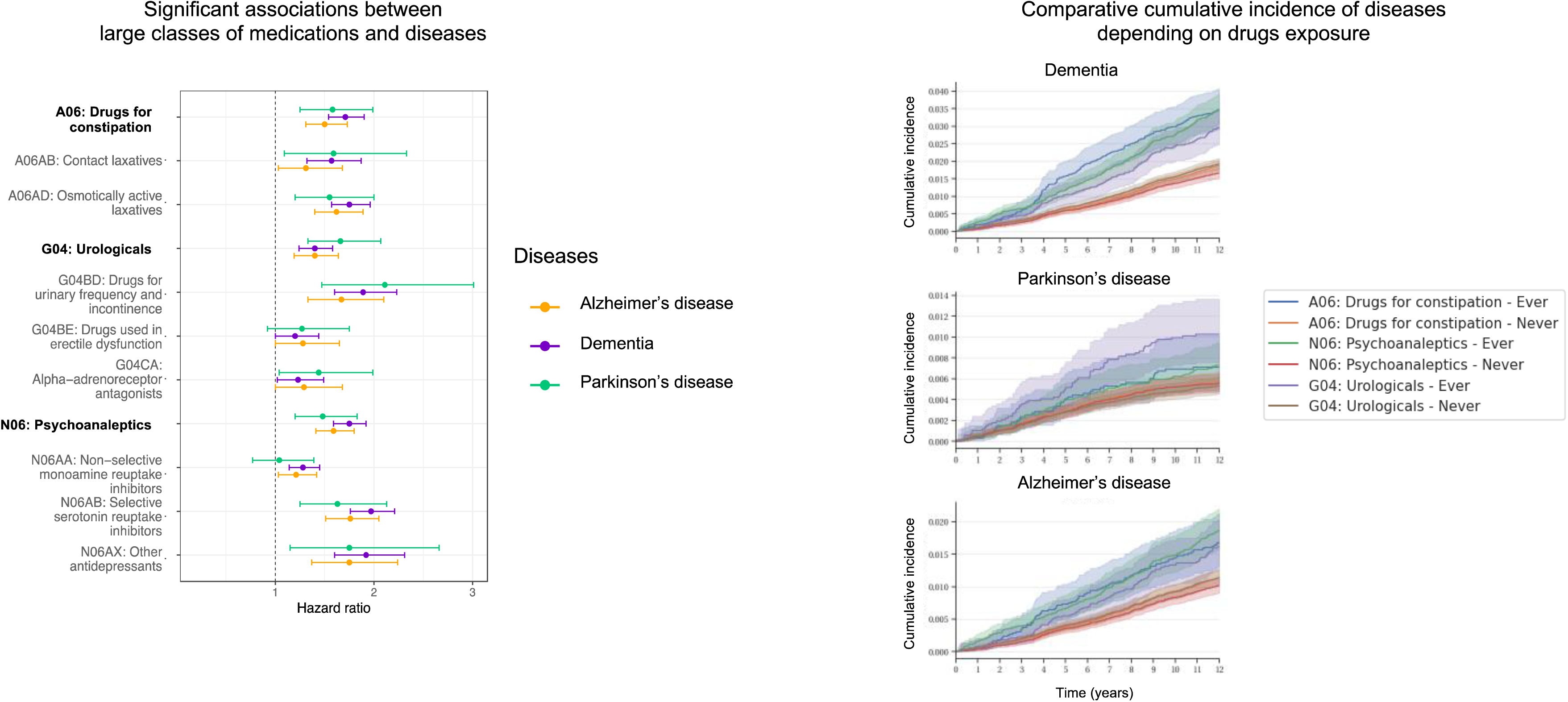
Feature selection based on associations of medication families with the onset of AD, PD and dementia Panel A: Classes of medications (depth 3) significantly associated with AD, PD and dementia are shown. For each class, we also show subfamilies of treatments. Bars correspond to 95% CIs after correction for multiple comparisons. Panel B: Fine Gray models with considering death as a competing risk adjusted for age class, sex, Charlson Index and body mass index with 95% confidence interval.

### 2.6 Prediction models – training and validation

We focused on predicting the first occurrence of each neurodegenerative disease over three periods, i.e., within 2, 5, and 10 years, using a logistic regression model (Figure 2) trained for each time period. We used the above-mentioned exposure time-window regardless of the prediction task (e.g. 2, 5, or 10 years). Patients were considered diseased if they experienced the specified outcome during the diagnostic period, namely between 2011, January 1^st^ and 2015, December 31^st^, when the prediction task was configured for a 5-year span (figure 1 and 2). Patients who died during the prediction period were regarded as non-cases of the neurodegenerative outcome in the prediction task.

#### Prediction phase

For the prediction phase, we investigated two primary models: i) a model that incorporates sex, BMI, Charlson Comorbidity Index (CCI), and the medications chosen in the feature selection stage, as exposure parameters; ii) a model that incorporates the same parameters as the first model, but also includes ten specific disease diagnoses corresponding to ICD10 codes of interest that have been previously demonstrated to be significantly positively associated with AD in a prior study (see supplementary materials for the exact list).^20^

#### Training and validation process

We defined the training set as comprising 75% of the UK data, with the remaining 25% serving as the UK test set, and used a separate external French test set. A 5-fold cross-validation was performed on the training set to select the optimal method for subsampling the majority class. We report several performance metrics, including the area under the receiver operating characteristic curve (AUROC), the detection rate for a fixed false positive rate of 5%, and the false positive rate for a fixed detection rate of 50%. As a sensitivity analysis, we excluded patients with Mild Cognitive Impairment (MCI) (codes provided in the supplementary materials) during the exposure period. We calculated 95% confidence intervals (95% CIs) using 1000 bootstrap replications from the test data. The survival and cmprsk libraries in R were used for the survival model, while the sklearn package in Python version 3.6 was employed for the logistic regression model and model evaluation.Role of the funding source:

The funder of the study had no role in study design, data collection, data analysis, data interpretation, or writing of the report.

## Results

Table 1 presents the age and sex distributions of all participants, patients with AD, PD, or dementia, and those who died. The study included 99,131 UK participants (45,062 born in 1940; 54,069 born in 1945), eFigure 1. During follow-up, 1,810 patients were diagnosed with AD, 660 with PD, 3,061 with dementia, and 10,030 died. Women slightly outnumbered men, especially among AD and dementia patients, while men predominated in PD and deceased patients. The average diagnosis year was 2015 for PD and 2017 for AD and dementia. Patients diagnosed after 2011 took more medications and had more healthcare visits, particularly consuming more laxatives, antidiabetics, lipid-modifying agents, antidepressants, and anti-dementia drugs compared to the overall population.

**Table 1:** Characteristics of patients with Azheimer’s disease, Parkinson’s disease and dementia in the UK

|  | tot. Pop | Alzheimer's disease | Parkinson's disease | Dementia | MCI | Death |
| --- | --- | --- | --- | --- | --- | --- |
| n | 54069 (100.0 %) | 607 (1.1 %) | 306 (0.6 %) | 1040 (1.9 %) | 1530 (2.8 %) | 4362 (8.1 %) |
| Female | 28203 (52.2%) | 371 (61.1 %) | 108 (35.3 %) | 578 (55.6 %) | 780 (51.0 %) | 1892 (43.4 %) |
| Male | 25866 (47.8 %) | 236 (38.9 %) | 198 (64.7 %) | 462 (44.4 %) | 750 (49.0 %) | 2470 (56.6 %) |
| Age at baseline (in 2010) | 65 | 65 | 65 | 65 | 65 | 65 |
| Follow-up time | 12.0 (12.0-12.0) | 12.0 (10.04-12.0) | 12.0 (10.77-12.0) | 12.0 (9.91-12.0) | 12.0 (11.73-12.0) | 6.45 (3.28-9.18) |
| Time to disease |  | 6.77 (3.83-9.08) | 4.76 (2.71-7.4) | 6.8 (3.86-9.08) | 4.44 (2.12-7.46) | 6.45 (3.28-9.18) |
| Avg. visits per year | 6.0 (3.33-10.33) | 7.0 (3.67-12.67) | 7.0 (3.67-11.67) | 7.67 (4.0-14.0) | 8.33 (4.67-14.0) | 8.67 (4.67-14.33) |
| Avg. number of drugs prescribed per year | 8.33 (3.67-16.67) | 10.33 (4.0-21.67) | 8.67 (4.33-19.0) | 12.33 (4.92-25.0) | 12.33 (5.67-24.67) | 14.33 (6.67-27.58) |
| Avg. number of unique drugs prescribed per year (molecule level) | 2.67 (1.67-4.67) | 3.0 (1.33-5.0) | 3.0 (1.67-5.0) | 3.33 (1.67-6.0) | 3.67 (2.0-6.67) | 4.0 (2.33-6.33) |
| Avg. number of unique drugs prescribed per year (ATC depth 3) | 2.33 (1.33-3.67) | 2.33 (1.33-3.67) | 2.33 (1.33-4.0) | 2.67 (1.33-4.33) | 3.0 (1.67-4.67) | 3.0 (1.67-4.67) |
| Laxatives [A06] | 6080 (11.2 %) | 96 (15.8 %) | 41 (13.4 %) | 198 (19.0 %) | 294.0 (19.2 %) | 798.0 (18.3 %) |
| Drugs used in Diabetes [A10] | 5175 (9.6 %) | 63 (10.4 %) | 35 (11.4 %) | 139 (13.4 %) | 199.0 (13.0 %) | 743.0 (17.0 %) |
| RAAS Agents [C09] | 17808 (32.9 %) | 186 (30.6 %) | 92 (30.1 %) | 361 (34.7 %) | 554.0 (36.2 %) | 1880.0 (43.1 %) |
| Lipid Modifying Agents [C10] | 22314 (41.3 %) | 264 (43.5 %) | 121 (39.5 %) | 509 (48.9 %) | 737.0 (48.2 %) | 2252.0 (51.6 %) |
| Psychoanaleptics [N06] | 11650 (21.5 %) | 203 (33.4 %) | 80 (26.1 %) | 378 (36.3 %) | 523.0 (34.2 %) | 1261.0 (28.9 %) |
| Anti-dementia drugs [N06D] | 16 (0.03 %) | 5 (0.8 %) | 1 (0.3 %) | 8 (0.8 %) | 1.0 (0.06 %) | 6.0 (0.14 %) |
| MCI between 2008 and 2010 | 360 (0.7 %) | 45 (7.4 %) | 12 (3.92 %) | 63 (6.06 %) | 0 (0.0 %) | 45 (1.03 %) |
| MCI anytime before 2010 | 775 (1.4 %) | 59 (9.7 %) | 14 (4.58 %) | 82 (7.88 %) | 0 (0.0 %) | 84 (1.93 %) |

In total, 43 drug classes were analyzed in the UK database. Only three drug classes—laxatives (A06), urological drugs (G04), and antidepressants (N06)—were significantly associated with all three outcomes (AD, PD, and dementia) in the multivariate Fine and Gray model (eTable 1, Figure 3, Panel B). Further analysis of drug subclasses revealed significant associations with the three diseases: contact laxatives (A06AB), osmotic laxatives (A06AD), drugs for urinary frequency and incontinence (G04BD), selective serotonin reuptake inhibitors (N06AB), and other antidepressants (N06AX) (Figure 3, Panel A).

The logistic regression model trained on UK patients born in 1945 showed the strongest discriminatory power for predicting dementia at 5 years (AUROC 0.67, 95% CI 0.64-0.69), followed by AD (0.66, 0.63-0.69) and PD (0.58, 0.53-0.61). Prediction improved over shorter time spans, with AUROCs of 0.74 (AD), 0.73 (dementia), and 0.56 (PD) for 2-year predictions. At a 5% false-positive rate, the algorithm detected 38.4%, 26.4%, and 18.3% of dementia cases and 39.6%, 26.0%, and 17.2% of AD cases over 2, 5, and 10 years, respectively. Adding health conditions did not enhance performance (Model 2 in eTable 5). The model performed better for women in AD and dementia predictions, and for men in PD predictions (Figure 4, Table 2, eTable6).

**Figure 4:**
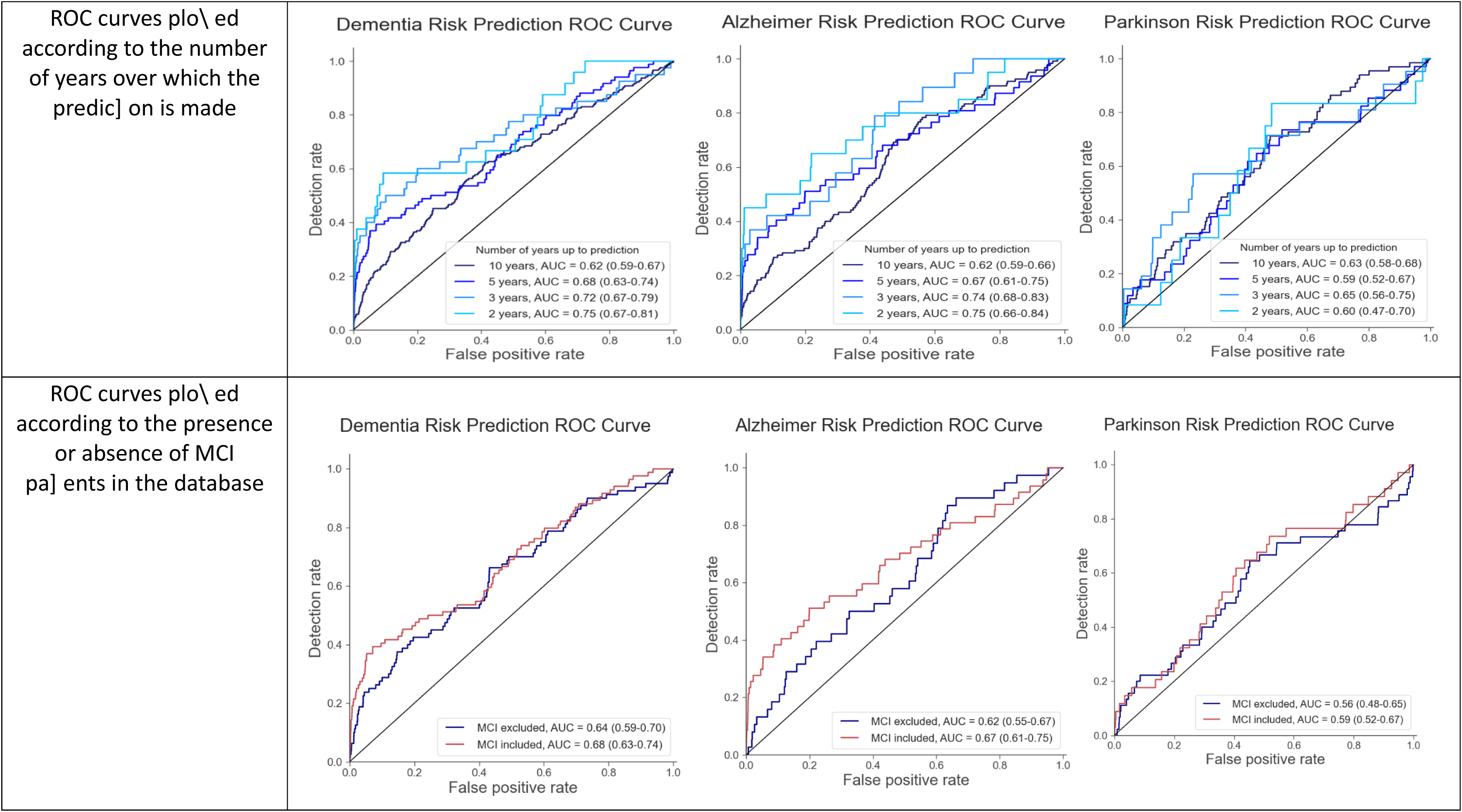
Change in Area Under Curve (AUC) for the prediction algorithms by duration of prediction and before and after the exclusion of patients diagnosed with MCI

**Table 2:** Performance of the predictive algorithms to estimate 2, 5 and 10-year disease risk for AD, PD and dementia at the age of 65 years Performance is analyzed using the model trained on the entire training population, with training, validation, and test Area Under the Curve (AUC) computed, alongside detection rates at a false positive rate of 5% and false positive rates at a detection rate of 50%.

| Disease |  | Prediction up to year | train AUC | test AUC | detection rate for a 5% fpr | fpr for a half detection rate | test AUC on FR |
| --- | --- | --- | --- | --- | --- | --- | --- |
| Dementia |  | 10 years | 0.65 (0.62-0.68) | 0.62 (0.61-0.63) | 18.3% (16.4%-20.0%) | 32.3% (30.0%-34.9%) | 0.55 (0.51-0.58) |
|  |  | 5 years | 0.71 (0.67-0.76) | 0.67 (0.64-0.69) | 26.4% (23.4%-28.6%) | 25.3% (21.4%-31.1%) | 0.62 (0.57-0.66) |
|  |  | 2 years | 0.80 (0.74-0.87) | 0.73 (0.68-0.77) | 38.4% (32.6%-44.2%) | 14.7% (8.9%-21.7%) | 0.68 (0.54-0.72) |
| Alzheimer |  | 10 years | 0.66 (0.62-0.69) | 0.61 (0.59-0.63) | 17.2% (14.8%-19.2%) | 35.5% (32.5%-38.3%) | 0.53 (0.47-0.58) |
|  |  | 5 years | 0.73 (0.68-0.78) | 0.66 (0.63-0.69) | 26.0% (22.7%-30.0%) | 27.8% (22.1%-33.7%) | 0.58 (0.51-0.62) |
|  |  | 2 years | 0.829 (0.74-0.92) | 0.74 (0.70-0.77) | 39.6% (31.2%-47.9%) | 13.8% (5.8%-21.6%) | 0.70 (0.55-0.77) |
| Parkinson |  | 10 years | 0.68 (0.64-0.72) | 0.61 (0.58-0.63) | 11.7% (8.6%-14.8%) | 35.6% (32.0%-39.5%) | 0.54 (0.50-0.58) |
|  |  | 5 years | 0.70 (0.64-0.74) | 0.58 (0.53-0.61) | 13.8% (8.9%-17.0%) | 37.4% (31.5%-46.7%) | 0.54 (0.48-0.60) |
|  |  | 2 years | 0.77 (0.68-0.85) | 0.56 (0.48-0.62) | 11.9% (4.6%-19.0%) | 37.9% (30.6%-50.2%) | 0.49 (0.28-0.68) |

Among UK patients, 1,587 were diagnosed with MCI between 2008 and 2010 (Table 1), with 10.8% later diagnosed with Alzheimer’s and 15.9% with dementia post-2011, rates 5 to 6 times higher than the overall prevalence (1.83% for Alzheimer’s and 3.09% for dementia). Excluding these MCI patients reduced the model’s performance: AUROCs for AD dropped from 0.67 (0.61-0.75) to 0.62 (0.55-0.67), for dementia from 0.68 (0.63-0.74) to 0.64 (0.59-0.70), and for PD from 0.59 (0.52-0.67) to 0.56 (0.48-0.65) (Figure 4.b, eTable 7).

## Discussion

Our study employs a methodology that integrates survival analysis and predictive modeling to investigate neurodegenerative disease outcomes using electronic health record (EHR) data. We found that we can reach high AUC for predicting dementias in the following 2 years, and to a lesser extent in the following 5 and 10 years, for individuals aged 65, using simple predictors. The strengths of this study lie in its utilization of routinely collected observational data, which allowed for the collection of a substantial number of exposures (ATC codes), not often feasible in randomized trials. Furthermore, by leveraging data from two distinct countries, we confirmed the robustness of our findings and enhanced the generalizability of our results.

Prevention strategies for neurodegenerative diseases are crucial, with early identification of at-risk individuals being a key component.^12^ However, conventional methods such as MRIs or PET scans for every individual over 65 are neither practical nor cost-effective. Various multifactorial risk prediction models have been developed to identify individuals at high risk of dementia, but their accuracy in same-aged populations remains limited.^6^ Recent findings indicate that existing models miss a significant portion of participants with incident dementia. Age alone shows equal promise as a predictor, highlighting the need for improvement in existing risk scores. One avenue for enhancing prediction models involves leveraging machine learning on electronic health records. Despite current limitations, machine learning algorithms offer a cost-effective means of predicting dementia risk from longitudinal clinical records.

Previous studies have attempted to predict dementia using electronic health records, but these efforts have been constrained to single countries and often rely heavily on age as a primary feature,^4,10,21,22^ posing challenges in interpretation. When analyzing models incorporating age as a predictive feature, reported classification performance across all ages may not offer a clear assessment of effectiveness due to inflated specificity from straightforwardly classifying younger patients as not at risk. When used in a same-aged group the AUC for the current risk scores (CAIDE, BDSI, ANU-ADRI) drop near 0.5.^6^ Additionally, while age often acts as a dominant predictor, other variables’ influences on predictions are typically more modest, when not included as cross-features with age, leading to an aggregated estimation of their impacts across all ages. In this perspective, we proposed a framework focused on predicting dementia onset at specific ages, notably at age 65, which aligns with the recommendation for preventive health check-ups in France. Our algorithm aims to serve as a pre-screening tool, identifying patients eligible for further diagnostic tests or interventions.

Our prediction algorithm demonstrates promising performance in detecting incident dementia, particularly when calibrated to achieve a 5% false-positive rate. Specifically, at this threshold, the algorithm detects 38% of incident dementia cases in a same-aged population at 2 years, 26% at 5 years and 18% at 10 years. This outperforms several widely used multifactorial 10-year risk prediction models, including the CAIDE score, CAIDE-APOE score, BDSI, and ANU-ADRI, which are to a large extent driven by age and detect only 9% to 16% of incident dementia cases, according to a recent study, even if all approaches have to be compared on the same patients before concluding to the superiority of a given model.^6^ Unlike those risk scores, our algorithm does not require clinical examination or responding to a questionnaire, highlighting its potential utility as a more cost-effective tool for identifying individuals at risk of dementia, thereby facilitating early intervention and management strategies.

Our analysis reveals heterogeneity in the predictive performance of the algorithm across different neurodegenerative diseases. Specifically, the algorithm demonstrates stronger discriminatory power for predicting dementia compared to AD and PD. This discrepancy may stem from differences in the underlying pathophysiology, risk factors, and disease progression trajectories among these conditions. Further research is warranted to elucidate these differences and refine prediction models tailored to each specific neurodegenerative disease. Sex-specific differences in the progression and diagnosis of neurodegenerative diseases may influence the predictive accuracy of machine learning models. Women exhibit distinct hormonal and genetic risk factors for AD and dementia, which may enhance model predictions in women.^23–25^ Conversely, PD presents differently in men, which may explain the better prediction accuracy for males due to unique clinical factors.^26^

The decision to employ a logistic regression model for our prediction analysis was informed by recent research demonstrating its efficacy in leveraging electronic health records for predictive modeling.^10^ Logistic regression allows also for straightforward interpretation of coefficients, facilitating the identification of significant predictors and their impact on disease risk. Its simplicity and transparency also makes it particularly appealing for clinical applications, where interpretability and ease of implementation are required.

Depression is recognized as a potential risk factor for neurodegenerative diseases.^27–29^ Previous studies have suggested a bidirectional relationship between depression and dementia, with depression increasing the risk of developing dementia and vice versa.^30^ Our findings align with this notion, as antidepressants (ATC code N06) which includes all anti-depressant drugs, showed significant associations with neurodegenerative disease outcomes. Furthermore, medications related to urinary incontinence (ATC code G04) and laxatives (ATC code A06) also displayed significant associations with neurodegenerative diseases. These associations might stem from the dual roles of these medications: facilitating the onset of neurodegenerative disease directly and/or administered to address prodromal symptoms of these diseases, which is the case for both anticholinergics and laxatives..^14,20^ For instance, anticholinergic medications commonly prescribed for incontinence management have been implicated in the acceleration of dementia onset.^31^ Prolonged use of these medications may disrupt cholinergic pathways in the brain, contributing to cognitive decline over time.^32,33^ Similarly, laxatives, frequently utilized to alleviate constipation, which is an established prodromal factor of neurodegenerative diseases, have also raised concerns regarding their potential role in neurodegenerative diseases.^34^

We observed sex differences in the predictive performance of the algorithm, with implications for risk stratification and intervention strategies. While the overall performance of the algorithm was robust, there were notable variations in prediction accuracy between male and female cohorts. These differences may reflect sex-specific risk factors, biological mechanisms, or healthcare-seeking behaviors that influence disease progression and detection.^25^ Understanding these disparities is essential for developing personalized approaches to dementia prevention and management that account for sex-specific considerations. Our analysis also revealed temporal variations in the predictive performance of the algorithm, suggesting dynamic changes in disease risk profiles over time. Specifically, the algorithm exhibited varying accuracy levels when predicting neurodegenerative disease onset over different time spans, such as 2-year vs 5-year versus 10-year horizons. These findings underscore the importance of considering temporal dynamics in risk prediction modeling^25^ and highlight the need for adaptable and responsive healthcare strategies that accommodate evolving disease trajectories and individual risk profiles.

Excluding patients diagnosed with MCI from the analysis had a noticeable impact on the algorithm’s predictive performance. MCI is often considered a transitional stage between normal aging and dementia, serving as an early indicator of neurodegenerative disease risk.^35^ Our findings suggest that incorporating MCI diagnoses improves prediction accuracy, emphasizing the importance of capturing subtle cognitive changes in early disease detection efforts. Further research into the predictive value of MCI and its implications for risk stratification is warranted to optimize dementia prevention and intervention strategies.^36^

The strengths of this study lie in its utilization of real-world observational data, which allowed for the collection of a substantial number of exposures (ATC codes), not often feasible in randomized trials. Furthermore, by leveraging data from two distinct countries, we improved the robustness of our findings, enhancing the generalizability of our results. Despite its strengths, our approach has certain limitations. The feature selection process of ATC codes carries the potential to introduce bias, given that it was executed on the entire UK dataset for computational efficiency. This procedure could lead to data leakage; however, this impact appears to have been mitigated as we were able to replicate the results in an independent study population. Additionally, the selection of medications based on Fine-Gray models might inadvertently overlook medications with potential relevance. Importantly, primary care data are characterized by two inherent challenges: the absence of key predictive variables and the reliability of ground truth. Primary care data inherently lacks crucial confounding variables such as education level, ethnicity, socio-economic status, and genetics, which could potentially introduce bias in hazard ratio estimations. Adding these variables could also help improve the performance of the different algorithms in the future. Additionally, it is imperative to acknowledge the potential for uncertainty in disease diagnoses made by general practitioners. It might result in a later diagnosis than in prospective cohorts in which each patient follows a protocol with planned visits with a neurologist.

## Conclusion

Leveraging machine learning on electronic health records presents a promising approach for the early identification of individuals at risk of neurodegenerative diseases. Our algorithm expands on the existing risk prediction models, reaching comparable predictive capacity and potentially providing a cost-effective and easily implementable solution for detecting incident dementia and other neurodegenerative diseases. We propose a new standard for training risk algorithms using electronic health records across multiple European countries and emphasize the importance of evaluating these algorithms without relying on age as a factor, particularly for screening purposes at specific ages.

## Supporting information

Supplemental results

## Data Availability

The data used in the preparation of this Article are available from the
Cegedim company upon reasonable request (info@the-health-
improvement-network.co.uk).

## Acknowledgements

The research leading to these results has received funding from the joint program in neurodegenerative diseases (JPND) ANR-21-JPW2-0002-01 (LeMeReND) and the program “Investissements d’avenir” ANR-10-IAIHU-06. This work was funded in part by the French government under management of Agence Nationale de la Recherche as part of the “Investissements d’avenir” program, reference ANR-19-P3IA-0001 (PRAIRIE 3IA Institute). PNS was supported by the Emil Aaltonen Foundation and the Finnish Medical Foundation. MK was supported by Wellcome Trust, UK (221854/Z/20/Z), National Institute on Aging (NIH), US (R01AG056477), Medical Research Council, UK (MR/R024227/1, MR/Y014154/1) and Academy of Finland (350426). BCD is supported by Inria, and a NHMRC CJ Martin fellowship (1161356).

## Ethical approval and consent statement

Ethical approval was obtained before carrying out this study (and for the use of datasets). The study was a retrospective analysis of secondary pseudo-anonymised patient data only and do not involve direct patient intervention. The study was approved by the THIN Scientific Research Committee (SRC; SRC reference 22-008-R2). There was no need for written informed consent from participants. No photographs, videos, or other information of recognizable persons is used in this article. Authorization for disclosure was, therefore, not necessary. For the UK database, data are only available to researchers carrying out approved medical research. Ethical approval was granted by the NHS South-East Multicentre Research Ethics Committee in 2003 (reference 03/01/073) for the establishment of the THIN database, and was updated in 2011. A further update was carried out and approval was granted in 2020 by the NHS South Central, Oxford C Research Ethics Committee (reference 20/SC/0011). For the French database, several audits were done by the Commission Nationale de l’Informatique et des Libertés, the French authority responsible for the protection of personal data. Data are available from GERS SAS for researchers who meet the criteria for access to confidential data.

## Contributors

KZ, CM, SD, MK and TN conceived and designed the study. YS and BL extracted the data from the THIN database. KZ,CM,MA,TN analyzed the data and ensured data quality. KZ and CM generated the figures. All authors interpreted the results and contributed to the writing of the final version of the manuscript.

## Data sharing

The data used in the preparation of this article are available from the Cegedim company upon reasonable request.

