## Supplemental results for "Machine learning prediction algorithms for 2- , 5- and 10-year risk of Alzheimer’s, Parkinson’s and dementia at age 65: a study using medical records from France and the UK General Practitioners"

eFigure 1: Selection of study participants in UK and FR Thin Data

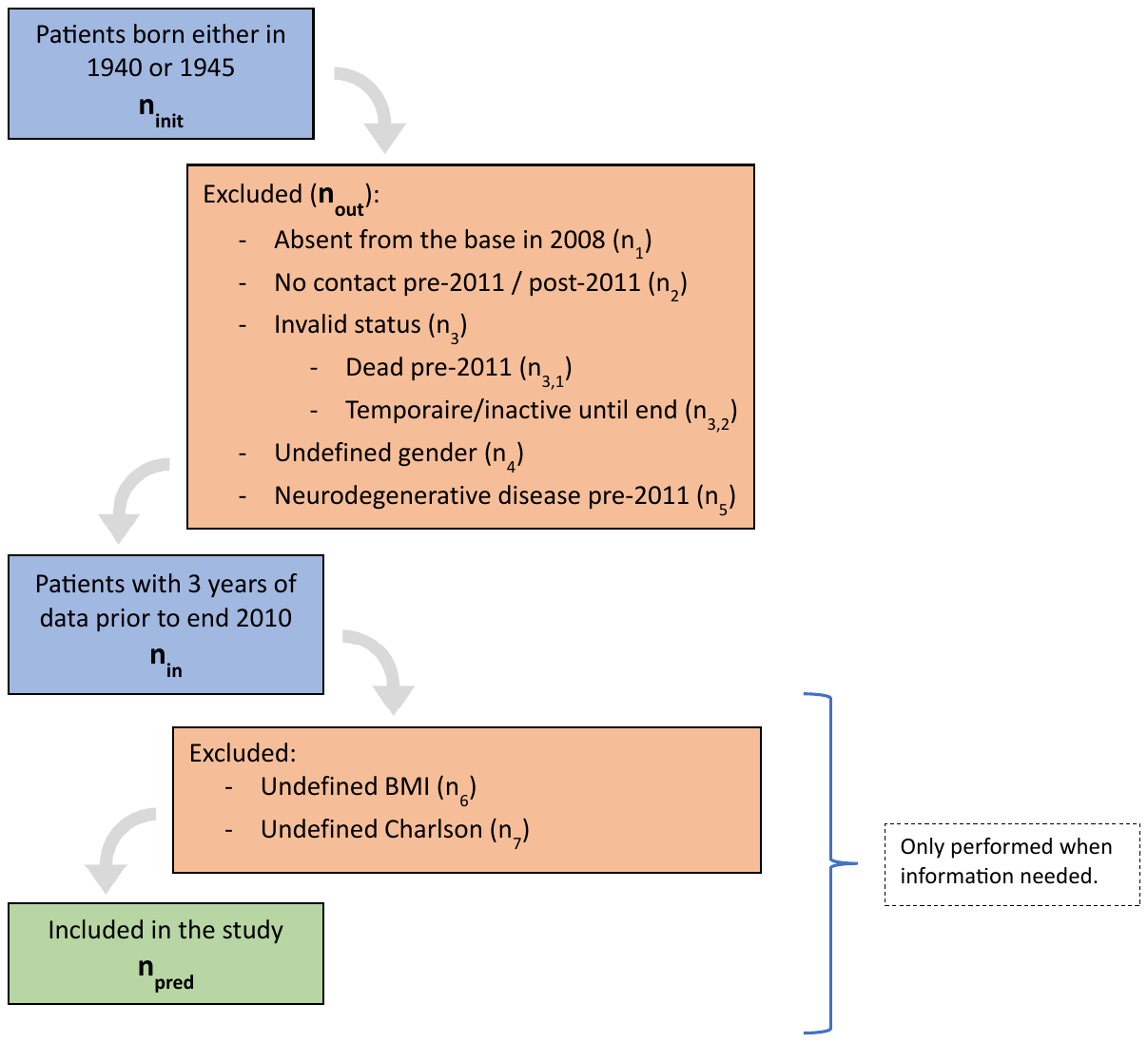

|  | **FR 65** | **UK 65** | **FR 70** | **UK 70** |
| --- | --- | --- | --- | --- |
| **n_init_** | **54,438** | **61,696** | **46,695** | **51,433** |
| **n_out_** | **32,080** | **7,627** | **28,235** | **6,371** |
| n_1_ | 28,172 | 3,592 | 25,095 | 2,695 |
| n_2_ | 3,736 | 3,605 | 2,886 | 3,016 |
| n_3_ | 0 | 41 | 1 | 35 |
| n_3,1_ | 0 | 11 | 0 | 18 |
| n_3,2_ | 0 | 30 | 1 | 17 |
| n_4_ | 0 | 2 | 0 | 0 |
| n_5_ | 172 | 387 | 253 | 625 |
| n_in_ | 22,358 | 54,069 | 18,460 | 45,062 |
| n_6_ | 9,243 | 2,681 | 7,716 | 1,950 |
| n_7_ | 446 | 528 | 0 | 458 |
| **n_pred_** | **13,115** | **50,899** | **10,744** | **42,686** |

Detailed supplementary methods:

Supplementary list of codes used to define outcomes :

AD was defined using the ICD-10 code F00 (Alzheimer’s dementia) and G30 (Alzheimer’s disease), PD using ICD-10 code G20 (Parkinson’s disease), and all-cause dementia with codes F00 (Alzheimer’s dementia), F01 (vascular dementia), F02 (Dementia in the course of other diseases classified elsewhere), F03 (Dementia, unspecified), F04 (Organic amnesic syndrome, not induced by alcohol or other psychoactive substances), F05.1 (Delirium added to dementia), G30 (Alzheimer’s disease), G31.0 (Circumscribed brain atrophy), G31.1 (Senile cerebral degeneration, not elsewhere classified), and G31.8 (Other specified degenerative conditions of the nervous system).

MCI ICD-10 codes: F06.7 (Mild Cognitive Impairment), R41.8 (Symptoms and signs related to cognitive functions and consciousness, other and unspecified), and R41 (Other symptoms related to cognitive functions and consciousness)

Precise computing of BMI and CCI

Exposure parameters encompass a variety of factors. Firstly, we calculated the Body Mass Index (BMI) of patients using recorded height and weight data. For each patient, the median height across all records and the mean weight over the exposure period were utilized. In cases where there were no records between 2008 and 2010, we used the weight recorded closest to the exposure period. Additionally, we computed a mean value of the Charlson Comorbidity Index (CCI) over the exposure period. CCI is an integer-based metric designed to quantify the risk of mortality at 1 and 10 years based on the analysis of 19 comorbidities in patients.^19^ Furthermore, we considered all prescriptions received during the exposure period, using the first 3 characters of the Anatomical Therapeutic Chemical (ATC) code, defining medication category. Similarly, we compiled all medical diagnoses received by the patient during the exposure period, extracting the ICD-10 codes in the same manner as for the outcome diseases.

List of diagnostic codes associated with Alzheimer’s disease :

ICD10 codes of interest that have been previously demonstrated to be significantly positively associated with AD in a prior study.^20^ These ten diagnoses included major depressive disorder [ICD-10 F32], anxiety [F41], reaction to severe stress and adjustment disorders [F43], hearing loss [H91], constipation [K59], spondylosis [M47], memory loss symptom [R41], malaise and fatigue [R53], syncope and collapse [R55], abnormal weight loss [R63].

eTable1: Association of each medication with each disease

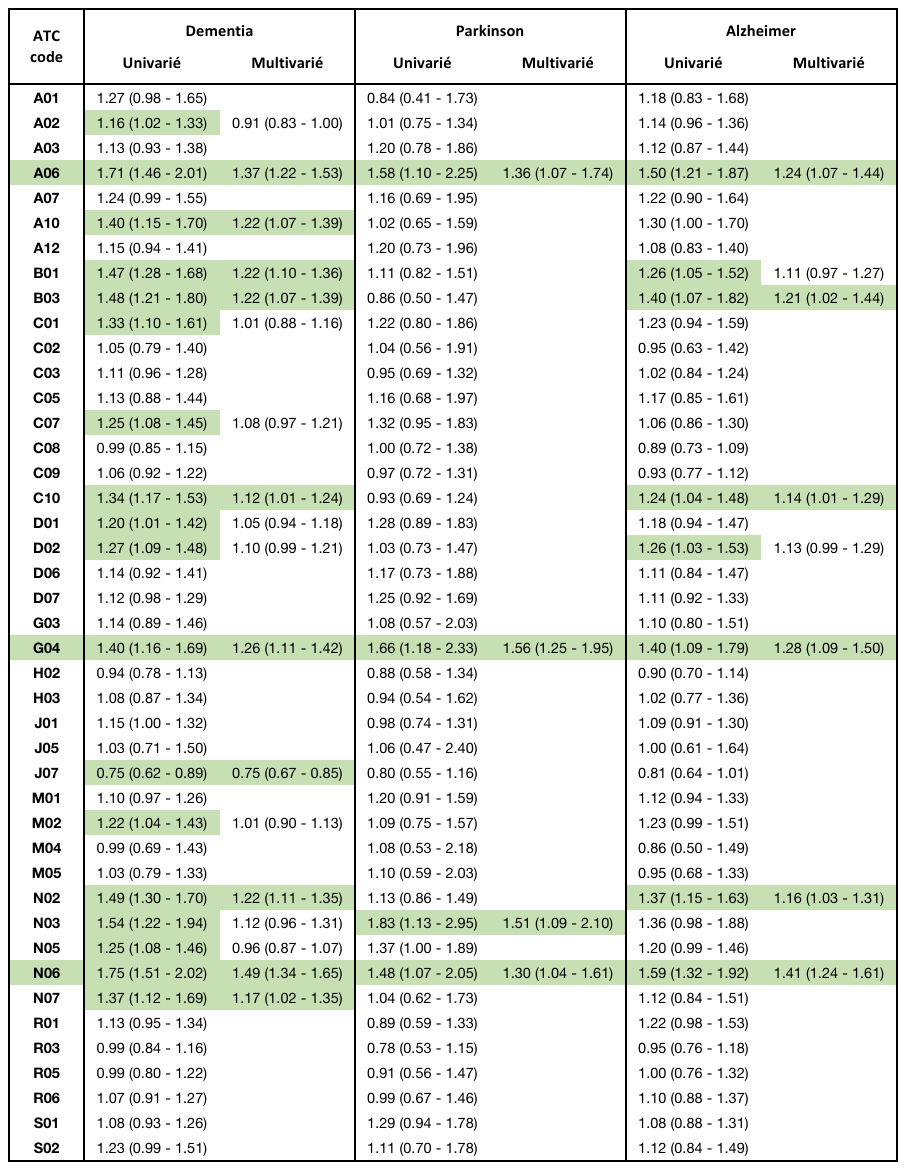

eTable 2: Characteristics of patients with outcome diseases, controls and deceased patients in full UK and FR database

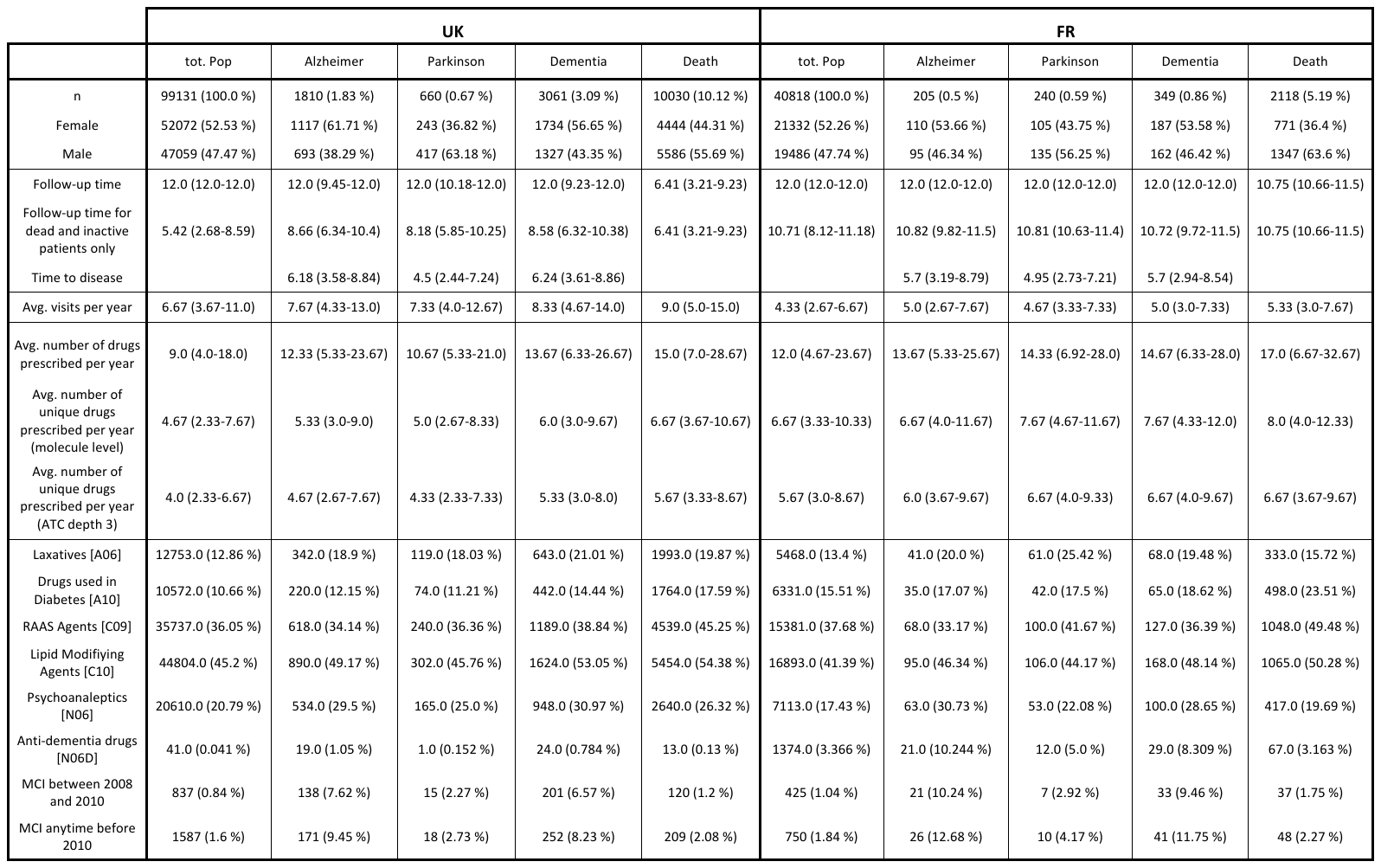

eTable 3: Characteristics of patients with outcome diseases, controls and deceased patients in UK 65 and 70 database

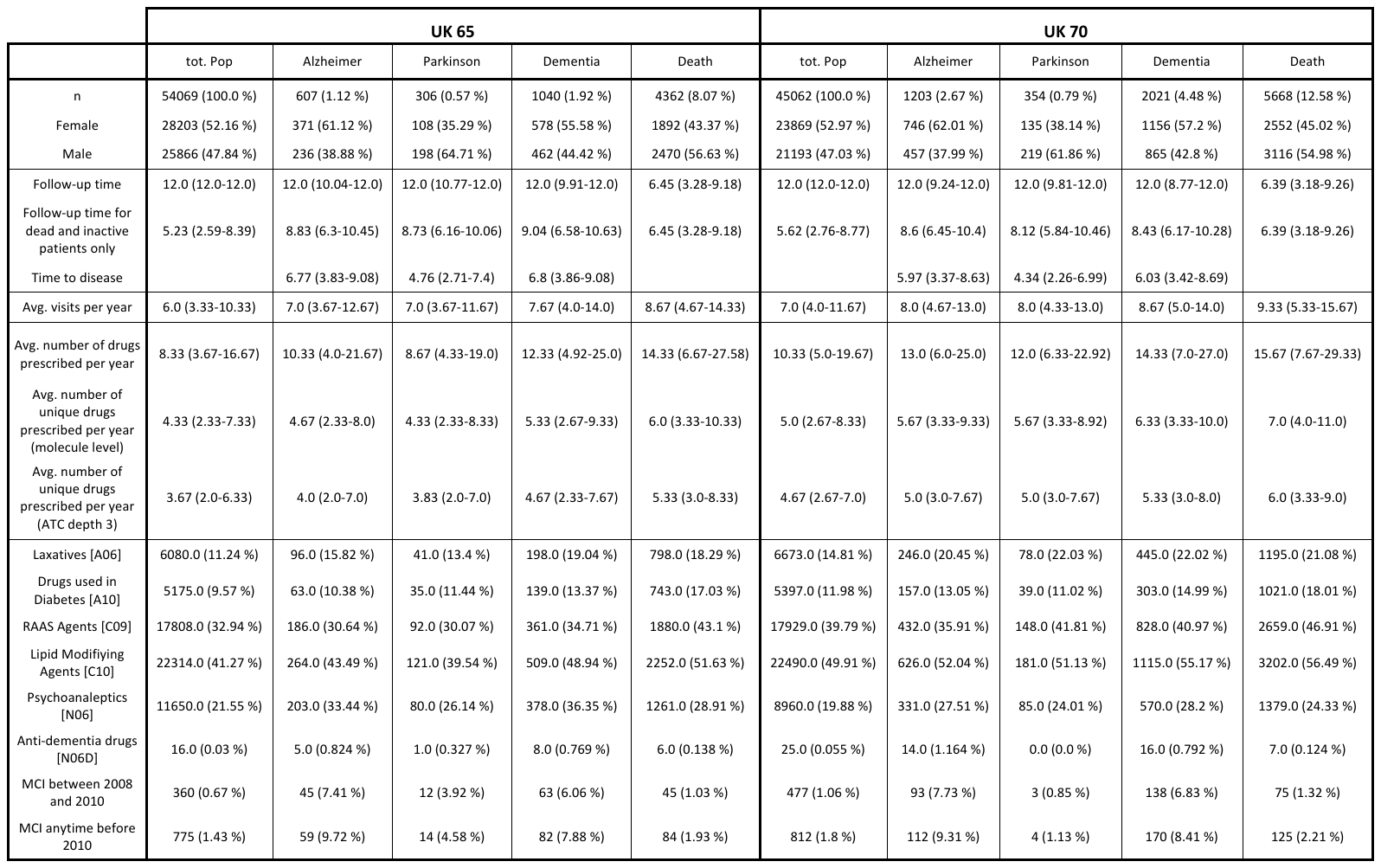

eTable 4: Characteristics of patients with outcome diseases, controls and deceased patients in FR 65 and 70 database

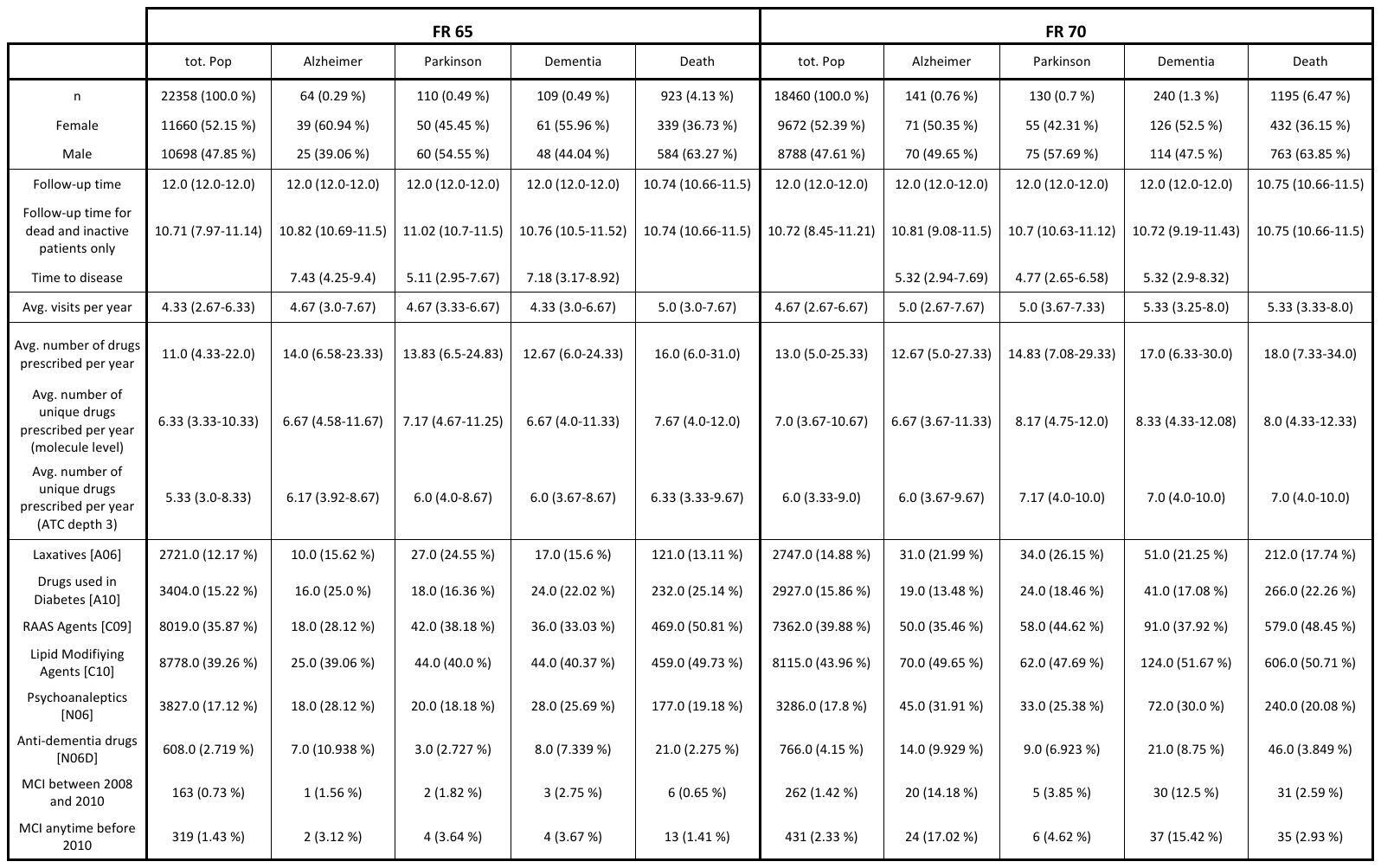

eTable 5: Performance of the classification algorithm to estimate disease risk for each outcome disease up to several numbers of years, for model 2

| **Disease** | **Prediction up to year** | **65 in 2010** | | | | | **70 in 2010** | | | | |
| --- | --- | --- | --- | --- | --- | --- | --- | --- | --- | --- | --- |
|  |  | **train AUC** | **test AUC** | **detection rate for a 5% fpr** | **fpr for a half detection rate** | **test AUC on FR** | **train AUC** | **test AUC** | **detection rate for a 5% fpr** | **fpr for a half detection rate** | **test AUC on FR** |
| **Dementia** | 10 years | 0.64 (0.628-0.65) | 0.624 (0.597-0.663) | 18.9% (15.1%-23.5%) | 31.9% (24.9%-35.7%) | 0.551 (0.539-0.564) | 0.62 (0.599-0.641) | 0.601 (0.586-0.609) | 14.5% (13.1%-15.5%) | 35.8% (34.3%-37.6%) | 0.591 (0.568-0.608) |
|  | 5 years | 0.696 (0.679-0.711) | 0.679 (0.634-0.733) | 28.1% (21.4%-35.2%) | 23.3% (13.7%-34.5%) | 0.627 (0.608-0.642) | 0.68 (0.651-0.708) | 0.643 (0.623-0.656) | 20.4% (18.8%-22.1%) | 29.7% (27.5%-32.5%) | 0.616 (0.57-0.644) |
|  | 3 years | 0.753 (0.736-0.769) | 0.723 (0.673-0.784) | 34.1% (25.6%-43.4%) | 15.6% (7.7%-30.1%) | 0.681 (0.667-0.698) | 0.707 (0.671-0.756) | 0.653 (0.628-0.674) | 25.9% (23.3%-28.2%) | 27.2% (23.5%-31.2%) | 0.616 (0.569-0.647) |
|  | 2 years | 0.778 (0.756-0.799) | 0.75 (0.673-0.81) | 38.7% (27.3%-50.0%) | 14.6% (4.6%-32.5%) | 0.693 (0.663-0.718) | 0.753 (0.71-0.792) | 0.694 (0.658-0.714) | 31.9% (27.6%-34.7%) | 19.2% (14.1%-25.2%) | 0.591 (0.536-0.635) |
| **Alzheimer** | 10 years | 0.643 (0.63-0.655) | 0.623 (0.592-0.66) | 17.4% (13.6%-23.6%) | 36.0% (28.2%-41.4%) | 0.524 (0.505-0.542) | 0.631 (0.607-0.655) | 0.596 (0.582-0.608) | 15.2% (13.6%-16.9%) | 36.7% (34.6%-39.1%) | 0.563 (0.529-0.592) |
|  | 5 years | 0.703 (0.68-0.724) | 0.672 (0.614-0.741) | 28.3% (20.0%-40.0%) | 26.2% (12.4%-40.3%) | 0.588 (0.56-0.612) | 0.677 (0.65-0.709) | 0.626 (0.61-0.648) | 22.2% (19.9%-24.2%) | 31.2% (27.7%-36.0%) | 0.572 (0.502-0.623) |
|  | 3 years | 0.771 (0.749-0.794) | 0.74 (0.684-0.824) | 41.5% (28.6%-57.7%) | 10.9% (3.2%-26.0%) | 0.708 (0.667-0.733) | 0.726 (0.678-0.782) | 0.656 (0.627-0.686) | 29.3% (26.6%-32.1%) | 25.6% (20.1%-31.4%) | 0.611 (0.554-0.66) |
|  | 2 years | 0.786 (0.763-0.823) | 0.751 (0.661-0.831) | 47.1% (30.0%-61.6%) | 7.0% (0.9%-24.2%) | 0.711 (0.641-0.765) | 0.772 (0.707-0.83) | 0.683 (0.638-0.707) | 34.5% (31.9%-38.8%) | 20.7% (15.1%-27.5%) | 0.571 (0.48-0.643) |
| **Parkinson** | 10 years | 0.654 (0.637-0.672) | 0.627 (0.585-0.673) | 12.3% (6.6%-19.6%) | 34.5% (28.7%-39.7%) | 0.551 (0.526-0.568) | 0.656 (0.615-0.699) | 0.586 (0.557-0.607) | 10.3% (8.0%-13.0%) | 37.5% (33.2%-43.3%) | 0.564 (0.536-0.591) |
|  | 5 years | 0.644 (0.621-0.672) | 0.592 (0.527-0.67) | 15.2% (6.8%-25.0%) | 36.7% (26.5%-44.8%) | 0.557 (0.519-0.588) | 0.694 (0.658-0.745) | 0.614 (0.575-0.64) | 14.5% (9.7%-18.6%) | 33.9% (28.1%-39.5%) | 0.582 (0.536-0.614) |
|  | 3 years | 0.707 (0.68-0.739) | 0.651 (0.562-0.741) | 19.2% (7.6%-33.3%) | 27.6% (18.8%-37.6%) | 0.431 (0.366-0.469) | 0.748 (0.7-0.811) | 0.651 (0.581-0.688) | 16.3% (8.7%-20.7%) | 28.3% (22.8%-36.8%) | 0.601 (0.532-0.675) |
|  | 2 years | 0.7 (0.667-0.739) | 0.598 (0.478-0.697) | 15.0% (0.0%-30.0%) | 33.0% (19.7%-50.3%) | 0.485 (0.376-0.682) | 0.797 (0.719-0.855) | 0.697 (0.612-0.739) | 16.7% (8.3%-25.0%) | 22.4% (15.8%-34.4%) | 0.574 (0.528-0.617) |

eTable 6: Performance of the classification algorithm to estimate 5-year disease risk for each outcome disease

| **Disease** | **Model** | **tested on** | **65 in 2010, UK** | | | | |  | **70 in 2010, UK** | | | | |
| --- | --- | --- | --- | --- | --- | --- | --- | --- | --- | --- | --- | --- | --- |
|  |  |  | **train AUC** | **test AUC** | **detection rate for a 5% fpr** | **fpr for a half detection rate** | **test AUC on FR** |  | **train AUC** | **test AUC** | **detection rate for a 5% fpr** | **fpr for a half detection rate** | **test AUC on FR** |
| **All dementias** | **1** | All | 0.653 (0.637-0.668) | 0.647 (0.605-0.695) | 17.8% (11.9%-25.3%) | 30.1% (20.8%-37.7%) | 0.636 (0.621-0.65) |  | 0.627 (0.616-0.636) | 0.618 (0.594-0.648) | 11.6% (9.0%-14.4%) | 31.7% (27.5%-35.7%) | 0.65 (0.645-0.655) |
|  |  | Only women | 0.664 (0.642-0.686) | 0.652 (0.591-0.71) | 16.5% (8.9%-25.1%) | 25.6% (16.0%-40.2%) | 0.524 (0.501-0.54) |  | 0.605 (0.592-0.619) | 0.6 (0.557-0.634) | 11.5% (7.8%-14.4%) | 35.3% (28.6%-43.2%) | 0.678 (0.672-0.684) |
|  |  | Only men | 0.639 (0.615-0.663) | 0.636 (0.574-0.704) | 18.9% (10.5%-27.9%) | 32.5% (20.6%-44.6%) | 0.756 (0.744-0.765) |  | 0.641 (0.623-0.658) | 0.638 (0.589-0.694) | 13.8% (9.7%-18.9%) | 26.8% (21.3%-36.2%) | 0.632 (0.627-0.636) |
|  | **2** | All | 0.696 (0.679-0.711) | 0.679 (0.634-0.733) | 28.1% (21.4%-35.2%) | 23.3% (13.7%-34.5%) | 0.627 (0.608-0.642) |  | 0.668 (0.657-0.679) | 0.655 (0.619-0.679) | 20.3% (16.7%-24.3%) | 27.8% (24.4%-33.1%) | 0.629 (0.618-0.64) |
|  |  | Only women | 0.707 (0.685-0.732) | 0.697 (0.633-0.753) | 28.7% (19.3%-40.0%) | 17.0% (9.9%-32.8%) | 0.517 (0.499-0.541) |  | 0.654 (0.641-0.67) | 0.641 (0.599-0.68) | 22.5% (18.0%-27.8%) | 30.3% (22.8%-38.9%) | 0.656 (0.639-0.67) |
|  |  | Only men | 0.682 (0.657-0.709) | 0.665 (0.593-0.747) | 25.0% (17.0%-33.3%) | 29.3% (13.8%-42.5%) | 0.748 (0.731-0.762) |  | 0.676 (0.659-0.693) | 0.663 (0.614-0.713) | 19.3% (13.0%-25.3%) | 23.9% (18.4%-31.8%) | 0.612 (0.598-0.624) |
| **Alzheimer** | **1** | All | 0.655 (0.635-0.675) | 0.643 (0.58-0.708) | 15.1% (8.3%-23.1%) | 31.3% (21.4%-41.7%) | 0.587 (0.553-0.611) |  | 0.615 (0.605-0.627) | 0.602 (0.565-0.626) | 9.9% (6.2%-14.1%) | 33.8% (29.1%-41.2%) | 0.599 (0.588-0.608) |
|  |  | Only women | 0.65 (0.623-0.678) | 0.629 (0.541-0.708) | 12.5% (5.4%-23.6%) | 31.1% (19.4%-46.9%) | 0.532 (0.492-0.567) |  | 0.593 (0.577-0.611) | 0.58 (0.532-0.629) | 9.3% (3.9%-14.1%) | 36.5% (28.2%-42.5%) | 0.616 (0.61-0.624) |
|  |  | Only men | 0.632 (0.606-0.663) | 0.626 (0.531-0.699) | 14.3% (5.3%-23.9%) | 33.3% (20.7%-47.9%) | 0.686 (0.665-0.706) |  | 0.621 (0.604-0.637) | 0.603 (0.537-0.666) | 8.2% (4.2%-14.0%) | 32.5% (21.7%-45.4%) | 0.61 (0.598-0.629) |
|  | **2** | All | 0.703 (0.68-0.724) | 0.672 (0.614-0.741) | 28.3% (20.0%-40.0%) | 26.2% (12.4%-40.3%) | 0.588 (0.56-0.612) |  | 0.663 (0.651-0.678) | 0.64 (0.595-0.672) | 22.4% (17.7%-28.1%) | 29.5% (23.7%-38.0%) | 0.585 (0.561-0.606) |
|  |  | Only women | 0.701 (0.674-0.73) | 0.672 (0.579-0.755) | 27.3% (17.2%-42.9%) | 25.2% (8.9%-43.8%) | 0.555 (0.527-0.584) |  | 0.656 (0.64-0.677) | 0.634 (0.58-0.688) | 25.8% (19.2%-33.4%) | 31.1% (19.7%-38.5%) | 0.6 (0.572-0.625) |
|  |  | Only men | 0.677 (0.641-0.715) | 0.652 (0.534-0.766) | 23.8% (11.1%-37.5%) | 32.6% (8.4%-47.6%) | 0.699 (0.652-0.727) |  | 0.655 (0.635-0.675) | 0.63 (0.547-0.693) | 19.5% (12.7%-26.7%) | 29.2% (18.8%-45.2%) | 0.597 (0.546-0.639) |
| **Parkinson** | **1** | All | 0.618 (0.596-0.641) | 0.59 (0.536-0.669) | 9.6% (2.3%-14.7%) | 37.1% (26.3%-45.1%) | 0.574 (0.553-0.602) |  | 0.649 (0.631-0.671) | 0.634 (0.581-0.698) | 16.7% (9.3%-23.7%) | 28.1% (19.8%-40.8%) | 0.588 (0.579-0.594) |
|  |  | Only women | 0.549 (0.503-0.609) | 0.502 (0.336-0.628) | 6.9% (0.0%-18.2%) | 46.8% (25.8%-71.4%) | 0.642 (0.551-0.678) |  | 0.604 (0.579-0.64) | 0.611 (0.512-0.687) | 13.0% (0.0%-25.0%) | 37.9% (21.4%-52.4%) | 0.556 (0.535-0.574) |
|  |  | Only men | 0.572 (0.536-0.61) | 0.521 (0.43-0.594) | 7.4% (0.0%-15.9%) | 45.2% (31.9%-61.1%) | 0.551 (0.455-0.668) |  | 0.657 (0.636-0.69) | 0.649 (0.557-0.718) | 14.3% (4.2%-23.1%) | 28.2% (16.8%-37.3%) | 0.597 (0.587-0.608) |
|  | **2** | All | 0.644 (0.621-0.672) | 0.592 (0.527-0.67) | 15.2% (6.8%-25.0%) | 36.7% (26.5%-44.8%) | 0.557 (0.519-0.588) |  | 0.667 (0.649-0.689) | 0.641 (0.571-0.695) | 17.0% (11.3%-26.5%) | 29.2% (18.7%-42.1%) | 0.585 (0.57-0.601) |
|  |  | Only women | 0.57 (0.521-0.634) | 0.492 (0.358-0.612) | 8.0% (0.0%-25.0%) | 52.1% (25.9%-75.0%) | 0.544 (0.432-0.626) |  | 0.625 (0.592-0.659) | 0.615 (0.487-0.703) | 12.5% (3.5%-22.3%) | 33.4% (20.3%-54.4%) | 0.549 (0.522-0.566) |
|  |  | Only men | 0.621 (0.585-0.651) | 0.551 (0.446-0.643) | 19.0% (8.0%-33.3%) | 44.3% (23.6%-62.9%) | 0.583 (0.47-0.667) |  | 0.682 (0.661-0.715) | 0.658 (0.565-0.731) | 16.3% (7.4%-28.2%) | 24.0% (15.3%-42.0%) | 0.635 (0.619-0.648) |

eTable 7: Performance of the classification algorithm to estimate 5-year disease risk for each outcome disease, depending on the inclusion of MCI patients

| **Disease** | **Patients with MCI included** | **Model** | **65 in 2010, UK** | | | | |  | **70 in 2010, UK** | | | | |
| --- | --- | --- | --- | --- | --- | --- | --- | --- | --- | --- | --- | --- | --- |
|  |  |  | **train AUC** | **test AUC** | **detection rate for a 5% fpr** | **fpr for a half detection rate** | **test AUC on FR** |  | **train AUC** | **test AUC** | **detection rate for a 5% fpr** | **fpr for a half detection rate** | **test AUC on FR** |
| **All dementias** | **No** | 1 | 0.638 (0.621-0.657) | 0.637 (0.576-0.687) | 18.0% (12.3%-24.3%) | 31.7% (18.9%-43.3%) | 0.637 (0.615-0.653) |  | 0.614 (0.604-0.625) | 0.609 (0.574-0.635) | 10.1% (6.6%-13.5%) | 33.4% (29.3%-39.1%) | 0.636 (0.627-0.643) |
|  |  | 2 | 0.656 (0.638-0.674) | 0.643 (0.59-0.696) | 18.8% (12.0%-25.6%) | 28.5% (17.4%-39.4%) | 0.622 (0.6-0.642) |  | 0.621 (0.613-0.63) | 0.604 (0.575-0.631) | 9.9% (6.1%-13.0%) | 34.5% (30.7%-39.3%) | 0.612 (0.597-0.625) |
|  | **Yes** | 1 | 0.653 (0.637-0.668) | 0.647 (0.605-0.695) | 17.8% (11.9%-25.3%) | 30.1% (20.8%-37.7%) | 0.636 (0.621-0.65) |  | 0.627 (0.616-0.636) | 0.618 (0.594-0.648) | 11.6% (9.0%-14.4%) | 31.7% (27.5%-35.7%) | 0.65 (0.645-0.655) |
|  |  | 2 | 0.696 (0.679-0.711) | 0.679 (0.634-0.733) | 28.1% (21.4%-35.2%) | 23.3% (13.7%-34.5%) | 0.627 (0.608-0.642) |  | 0.668 (0.657-0.679) | 0.655 (0.619-0.679) | 20.3% (16.7%-24.3%) | 27.8% (24.4%-33.1%) | 0.629 (0.618-0.64) |
| **Alzheimer** | **No** | 1 | 0.639 (0.613-0.662) | 0.627 (0.561-0.685) | 14.3% (5.4%-22.0%) | 33.1% (20.6%-44.7%) | 0.584 (0.548-0.62) |  | 0.603 (0.588-0.615) | 0.585 (0.536-0.629) | 8.6% (4.8%-12.5%) | 36.0% (29.0%-44.5%) | 0.598 (0.58-0.612) |
|  |  | 2 | 0.648 (0.626-0.671) | 0.618 (0.557-0.669) | 14.1% (8.8%-23.1%) | 33.9% (21.1%-46.2%) | 0.581 (0.558-0.612) |  | 0.61 (0.591-0.622) | 0.573 (0.536-0.612) | 9.4% (5.3%-14.1%) | 38.0% (31.5%-44.7%) | 0.563 (0.529-0.596) |
|  | **Yes** | 1 | 0.655 (0.635-0.675) | 0.643 (0.58-0.708) | 15.1% (8.3%-23.1%) | 31.3% (21.4%-41.7%) | 0.587 (0.553-0.611) |  | 0.615 (0.605-0.627) | 0.602 (0.565-0.626) | 9.9% (6.2%-14.1%) | 33.8% (29.1%-41.2%) | 0.599 (0.588-0.608) |
|  |  | 2 | 0.703 (0.68-0.724) | 0.672 (0.614-0.741) | 28.3% (20.0%-40.0%) | 26.2% (12.4%-40.3%) | 0.588 (0.56-0.612) |  | 0.663 (0.651-0.678) | 0.64 (0.595-0.672) | 22.4% (17.7%-28.1%) | 29.5% (23.7%-38.0%) | 0.585 (0.561-0.606) |
| **Parkinson** | **No** | 1 | 0.605 (0.583-0.629) | 0.57 (0.498-0.64) | 6.7% (0.0%-13.3%) | 40.6% (28.1%-49.4%) | 0.555 (0.521-0.596) |  | 0.655 (0.636-0.674) | 0.633 (0.578-0.692) | 15.5% (10.2%-25.0%) | 29.5% (19.6%-42.8%) | 0.587 (0.574-0.595) |
|  |  | 2 | 0.633 (0.608-0.662) | 0.56 (0.488-0.642) | 9.5% (2.8%-16.1%) | 39.9% (29.2%-50.4%) | 0.549 (0.503-0.586) |  | 0.671 (0.654-0.688) | 0.635 (0.574-0.682) | 16.0% (9.8%-24.3%) | 31.3% (19.4%-43.4%) | 0.587 (0.575-0.601) |
|  | **Yes** | 1 | 0.618 (0.596-0.641) | 0.59 (0.536-0.669) | 9.6% (2.3%-14.7%) | 37.1% (26.3%-45.1%) | 0.574 (0.553-0.602) |  | 0.649 (0.631-0.671) | 0.634 (0.581-0.698) | 16.7% (9.3%-23.7%) | 28.1% (19.8%-40.8%) | 0.588 (0.579-0.594) |
|  |  | 2 | 0.644 (0.621-0.672) | 0.592 (0.527-0.67) | 15.2% (6.8%-25.0%) | 36.7% (26.5%-44.8%) | 0.557 (0.519-0.588) |  | 0.667 (0.649-0.689) | 0.641 (0.571-0.695) | 17.0% (11.3%-26.5%) | 29.2% (18.7%-42.1%) | 0.585 (0.57-0.601) |
